# The Impact of AI on Scientific Literature: A Surge in AI-Associated Words in Academic and Biomedical Writing

**DOI:** 10.1101/2024.05.31.24308296

**Authors:** Gwinyai Masukume

## Abstract

**Introduction:** The debut of ChatGPT on 30 November 2022, marked a new era in artificial intelligence (AI) innovation, ushering transformative progress in science and research efficiency. Yet, AI’s exact influence on scientific and biomedical literature remains unclear. Words like “delve”, “realm” and “underscore” are linked with AI chatbots rather than human authors, potentially signalling AI utilisation.

**Methods:** An extensive search of the electronic databases PubMed and Scopus was conducted, spanning from their inception to 28 May 2024, covering a period of 175 years from 1849 to 2024. The search identified occurrences of the words “delve”, “realm” and “underscore” individually and in various combinations within the scientific literature.

**Results:** Between 1849 and 2024, the simultaneous occurrence of “delve”, “realm” and “underscore” was observed exclusively during 2023 and 2024. Notably, in 2023 and 2024, “delve” and “underscore”, appeared together in 1,284 out of 1,299 (98.8%) PubMed articles (p-value = 0.0002) and 546 out of 566 (96.5%) Scopus articles (p-value= 0.0006) where they were used since 1849. The co-usage of these words increased by up to 85-fold in 2023 and 2024 compared to 2022 and prior years.

Additionally, among articles since 1849, in 2023 and 2024, “delve” appeared in 7,714 out of 17,537 (44.0%) PubMed and 6,360 out of 13,707 (46.4%) Scopus articles, “realm” in 3,825 out of 16,288 (23.5%) PubMed and 16,360 out of 82,714 (19.8%) Scopus articles, and “underscore” in 27,653 out of 95,606 (28.9%) PubMed and 39,154 out of 111,803 (35.0%) Scopus articles.

**Conclusion:** The analysis reveals a surge in the use of “delve”, “realm” and “underscore”—words presently associated with AI chatbots—during 2023 and 2024, highlighting the growing influence of AI chatbots like ChatGPT, introduced in November 2022, on academic writing. This trend highlights the increasing integration of AI-generated text in scientific articles, prompting further exploration into broader implications on research practices and scholarly communication.

## Introduction

The advent of ChatGPT on 30 November 2022, marked a watershed moment in artificial intelligence (AI), catalysing unprecedented advancements in scientific research efficiency [1]. However, amidst this revolution, the precise implications of AI on scholarly literature remain enigmatic [2]. Notably, certain words such as “delve”, “realm” and “underscore” have begun to be associated more prominently with AI chatbots rather than human authors [3], suggesting a subtle yet potentially transformative integration of AI into academic writing practices. AI’s promise in science and research is vast, but in fields like medicine and healthcare, caution is warranted due to potential confabulation and inaccuracy [4]. Therefore, the objective was to identify occurrences of the words “delve”, “realm” and “underscore” both independently and in various combinations within the corpus of scientific and biomedical literature [5].

## Methods

The methodological approach encompassed a comprehensive search of two widely recognised electronic databases, PubMed and Scopus. Spanning from their inception to 28 May 2024, the search covered a temporal period of 175 years, stretching from 1849 to 2024. Through this systematic analysis, the aim was to elucidate temporal trends and patterns indicative of AI’s evolving impact on academic writing practices.

The PubMed search query utilised the terms; “delve*” AND “realm*” AND “underscor*”. The wildcard (*) was employed to encompass variations such as delves, delved and delving. Boolean operators OR and AND were employed to generate various combinations of these words. A comparable methodology was adopted for the Scopus search, for instance, (TITLE-ABS-KEY(“delve” AND “realm” AND “underscore”)). For the full search syntax and search results, please see the Appendix.

Stata version 17BE was utilised for statistical analysis. A test for trend was conducted, with a p-value < 0.05 deemed statistically significant.

## Results

The comprehensive analysis of scientific literature spanning over a century unveiled intriguing patterns in the utilization of AI-associated words, particularly during the pivotal years of 2023 and 2024. Notably, the simultaneous occurrence of “delve”, “realm” and “underscore” was strikingly prevalent exclusively during this biennium, permeating 110 articles indexed in PubMed and 66 in Scopus.

Remarkably, in 2023 and 2024, “delve” and “underscore” appeared together in 1,284 out of 1,299 PubMed articles (98.8%) – Figure 1 – (p-value = 0.0002) and in 546 out of 566 Scopus articles (96.5%) – Figure 2 – (p-value = 0.0006) where they have been used since 1849. The frequency of these words being used together saw an increase of up to 85-fold in 2023 and 2024 compared to the period up to and including 2022.

**Figure 1.**
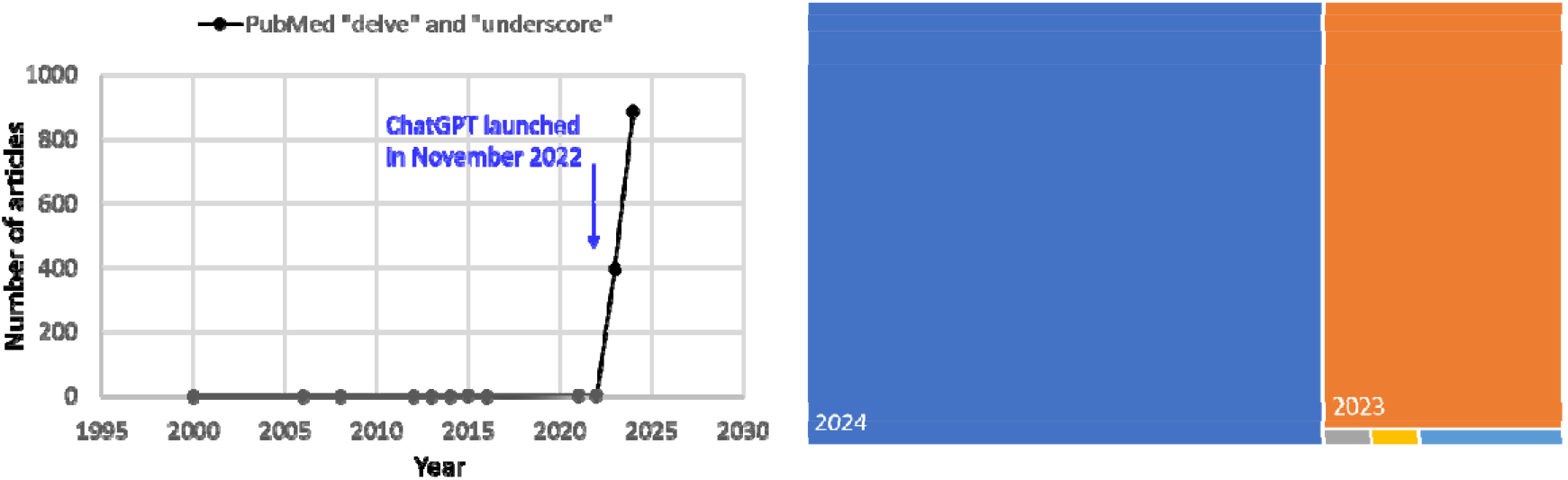
Simultaneous occurrence of “delve” and “underscore” in PubMed articles. This figure shows that “delve” and “underscore” appeared together in 1,284 out of 1,299 PubMed articles (98.8%) during 2023-2024, among articles they co-appeared in from 1849 to 2024, highlighting a significant increase in their co-occurrence during this two-year period.

**Figure 2.**
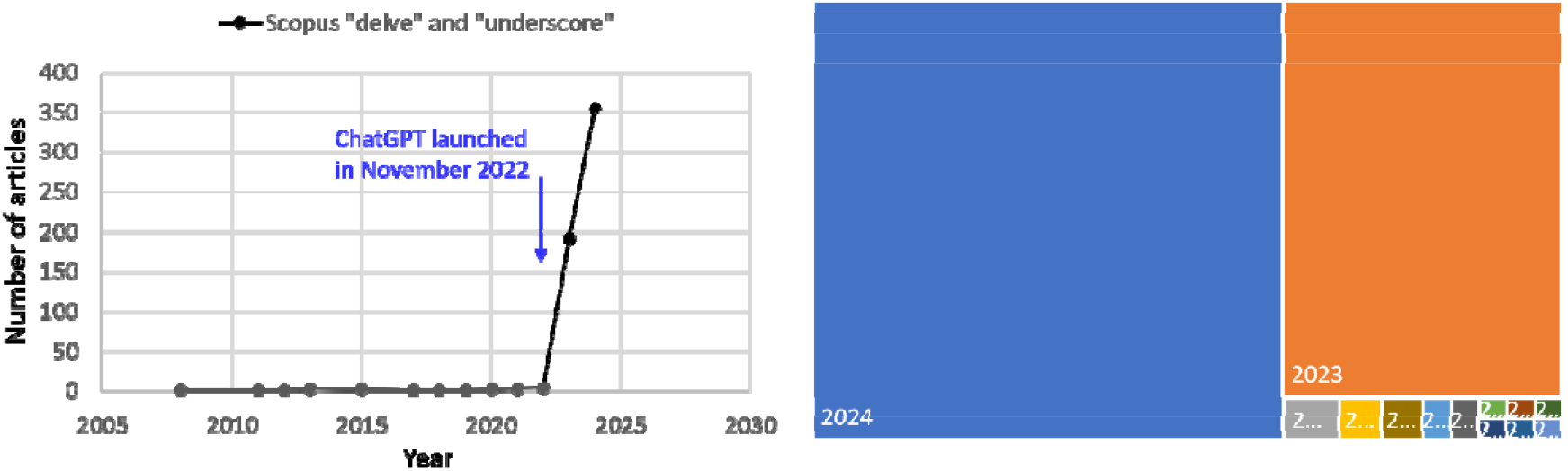
Simultaneous occurrence of “delve” and “underscore” in Scopus articles. This figure shows that “delve” and “underscore” appeared together in 546 out of 566 Scopus articles (96.5%) during 2023-2024, among articles they co-appeared in from 1849 to 2024, highlighting a significant increase in their co-occurrence during this two-year period.

Furthermore, among articles published since 1849, in 2023 and 2024, “delve” was found in 7,714 out of 17,537 PubMed articles (44.0%) – Figure 3 – and in 6,360 out of 13,707 Scopus articles (46.4%) – Figure 4. The word “realm” appeared in 3,825 out of 16,288 PubMed articles (23.5%) and in 16,360 out of 82,714 Scopus articles (19.8%). The word “underscore” was present in 27,653 out of 95,606 PubMed articles (28.9%) and in 39,154 out of 111,803 Scopus articles (35.0%).

**Figure 3.**
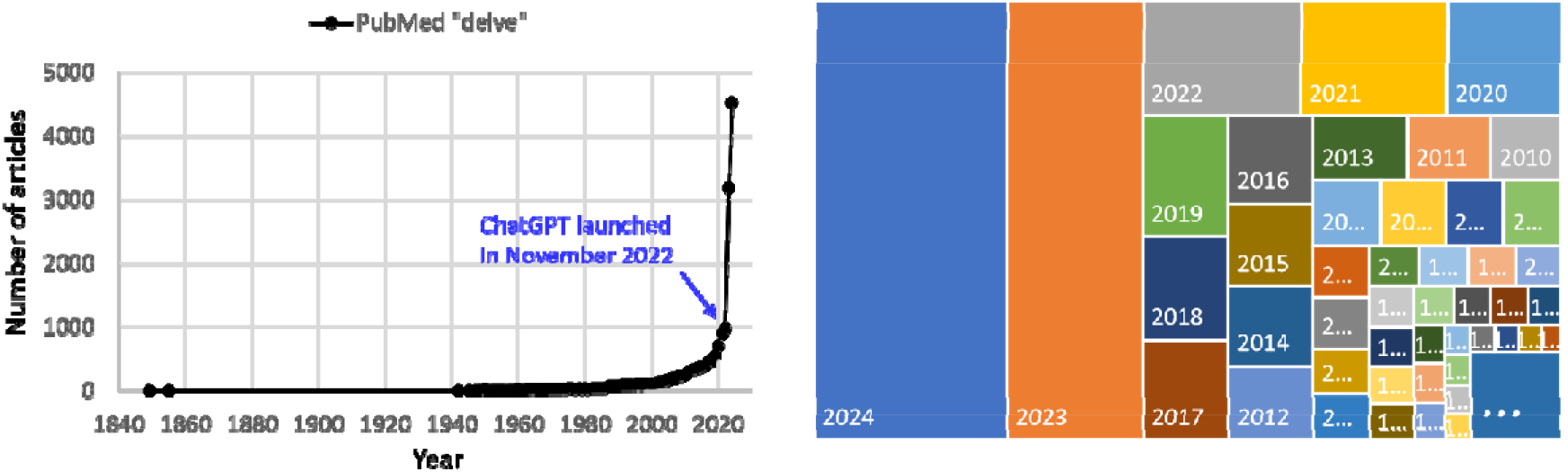
Occurrence of “delve” in PubMed. This figure illustrates the occurrence of the word “delve” in PubMed articles since 1849. Among PubMed articles where “delve” appears, it appeared in 44.0% (7,714 out of 17,537) during 2023 and 2024.

**Figure 4.**
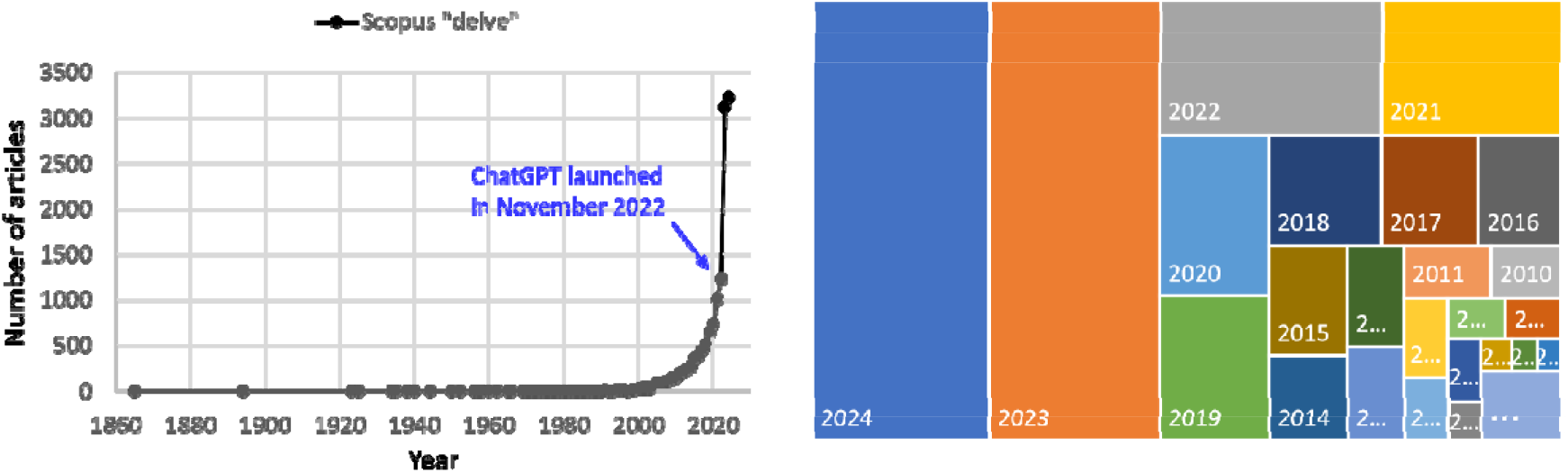
Occurrence of “delve” in Scopus. This figure illustrates the occurrence of the word “delve” in articles indexed in Scopus since 1849. Among Scopus articles where “delve” appears, it appeared in 46.4% (6,360 out of 13,707) during 2023 and 2024.

Between 2023 and 2024, at least one of these three words appeared in 36,983 out of 127,086 PubMed articles (29.1%) and in 59,608 out of 205,654 Scopus articles (29.0%) among those published since 1849 where at least one of these words was used.

## Discussion

This research letter reveals a significant shift in scientific discourse coinciding with the rise of AI tools like ChatGPT [6]. The pronounced surge in the usage of “delve”, “realm” and “underscore” during 2023 and 2024 in tens of thousands of scientific articles highlights the deep integration of AI-generated text in academic writing. The exclusive simultaneous occurrence of these words only during this period is striking, indicating a concentrated shift in academic writing practices. The co-occurrence of “delve” and “underscore” during 2023 and 2024 in PubMed and Scopus articles, with rates of 98.8% and 96.5% respectively among articles where they co-occur since 1849, emphasises this phenomenon. This stark departure from historical patterns highlights AI’s substantial impact on shaping scholarly discourse [3].

While AI enhances research efficiency [1], caution is warranted, particularly in fields like medicine and healthcare, where concerns about confabulation and factual inaccuracies have been raised [4]. The risk of homogenising academic writing, potentially masking individual researchers’ voices, necessitates careful consideration.

This study provides valuable insights into the evolving landscape of academic writing in the AI era. It further illuminates the growing integration of AI-generated text in scientific literature [5], highlighting both its promises and potential pitfalls. While valuable, the study’s reliance on publicly available databases and focus on specific keywords may introduce bias and overlook nuanced discourse, potentially limiting the breadth of its findings on AI integration in scholarly writing.

In conclusion, while AI offers unprecedented opportunities for research efficiency, its adoption in scholarly communication requires thoughtful management. Balancing the benefits of AI with the need to preserve the authenticity and accuracy of scientific and biomedical literature is essential for the future of academic research [2]. Further exploration of AI’s impact on academic writing practices is necessary to inform future research and publishing guidelines. The continuous examination of AI’s evolving role is crucial for ensuring the integrity and advancement of scholarly communication.

## Data Availability

All data produced in the present work are contained in the manuscript

## Data availability

Publicly available data was used.

## Ethics

Ethical approval was not required as the study was based on publicly available scholarly database search results.

